# Proteomic Signatures of Protected *APOE*-ε4 Carriers Reveal Causal Pathways Associated with Delayed Alzheimer’s Disease Onset

**DOI:** 10.64898/2026.04.30.26352191

**Authors:** Yann Le Guen, Junyoung Park, Andrés Peña-Tauber, Michael D. Greicius

**Author notes:** Corresponding author: Yann Le Guen Quantitative Sciences Unit 3180 Porter Drive, Palo Alto, CA 94304.

## Abstract

**INTRODUCTION:** *APOE*-ε4 is the strongest common genetic risk factor for Alzheimer’s disease (AD), yet many carriers remain cognitively unimpaired into late life. We tested whether a protected-ε4-first proteomic approach could identify plasma proteins associated with delayed clinical onset among ε4 carriers.

**METHODS:** We analyzed harmonized plasma proteomics from the Global Neurodegeneration Proteomics Consortium. Protected ε4 carriers (ε3/ε4 aged ≥75 years; ε4/ε4 aged ≥65 years; CDR=0; n=456) were compared with ε4 carriers with AD (n=1,096). Protein-wise linear models adjusted for age, sex, ε4 dosage, and plasma proteomic principal components. Top signals were integrated with high-confidence loss-of-function burden testing and plasma/CSF Mendelian randomization.

**RESULTS:** ε4 protected was associated with 721 protein levels. Integrated analyses prioritized proteins linked to ε4-modified disease biology, including LILRA5, DBI, BPNT1, PTEN, EPHA1, and PCDH10, and proteins aligned with broader AD-related change, including OMG, SELENOW, VAT1, and TPPP3. TREM2 and ACE were also identified, providing internal biological validation of the approach.

**DISCUSSION:** A protected-ε4-first plasma proteomic strategy highlights immune, synaptic, metabolic-stress, and myelin/axonal pathways that may delay AD onset and helps prioritize candidate ε4-specific modifiers for prevention-focused therapeutics.

## Background

Alzheimer’s disease (AD) is the most common cause of dementia and a major global public health challenge. Recent estimates indicate that tens of millions of people worldwide are living with dementia, and the number of individuals across the AD continuum is expected to rise sharply with population aging [1,2]. Contemporary biological frameworks define AD by its underlying pathologic processes rather than by symptoms alone, emphasizing that amyloid-β deposition, tau pathology, and neurodegeneration accumulate over a prolonged preclinical period before overt dementia emerges [3,4]. This extended interval between biological onset and clinical diagnosis creates an important opportunity for prevention-oriented discovery, particularly if mechanisms that delay symptom onset can be identified in living humans.

Genetic risk for AD is substantial, but not deterministic. The *APOE*-ε4 allele remains the strongest common genetic risk factor for late-onset AD, with dose-dependent effects on risk and age at onset [5,6]. However, *APOE*-ε4 is neither necessary nor sufficient for AD: many ε4 carriers never develop dementia, and estimated absolute risks vary by age, sex and ancestry [5]. Studies of *APOE* ε4 carriers therefore provide a particularly informative setting in which to search for protective factors that modify the relationship between inherited risk and clinical outcome [7–9].

Conceptually, this strategy draws on the literature distinguishing resistance from resilience in AD. Resistance refers to lower-than-expected pathology given risk, whereas resilience refers to better-than-expected cognitive or clinical function despite pathology [7]. In a research setting where biomarker is unavailable and subjects cannot be classified as either resistant or resilient, we favor the more agnostic term “protected”. In many observational cohorts, however, complete amyloid and tau biomarker data are unavailable, making delayed clinical onset among genetically high-risk individuals a pragmatic phenotype for enrichment of protective biology. In the *APOE*-ε4 context, defining cognitively unimpaired older ε4 carriers as resilient therefore provides an operational framework to identify biological factors associated with delayed dementia onset [7,8].

Blood-based proteomics offers a scalable strategy for discovering such biology in living humans. Large affinity-based platforms, including aptamer-based assays such as SomaScan, can quantify thousands of plasma proteins simultaneously and have shown suitable reproducibility for population-scale biomarker discovery studies [10,11]. These technologies enable systems-level analyses in cohorts that are substantially larger than those typically available for cerebrospinal fluid or neuroimaging studies [10,12,13]. The Global Neurodegeneration Proteomics Consortium (GNPC) recently harmonized multi-cohort plasma proteomic data at unprecedented scale, improving power for biomarker and target discovery across neurodegenerative diseases and aging [12]. However, an important limitation of conventional case–control proteomic analyses is that many proteins associated with AD diagnosis or cognitive decline may reflect downstream responses to neurodegeneration, inflammation, frailty, medication, or comorbidity rather than upstream modifiers of disease onset [7,12,13].

Human genetics can help address this limitation through causal triangulation. Large pQTL resources have now mapped genetic regulators of plasma and cerebrospinal fluid protein levels, providing instruments for causal inference and target prioritization [14–16]. Two-sample Mendelian randomization using pQTLs can strengthen inference regarding whether altered protein abundance is more likely to lie upstream of disease risk, particularly when combined with complementary evidence from colocalization and disease genetics [15–17]. In parallel, rare-variant gene-based analyses in large sequencing cohorts can identify genes in which predicted loss-of-function variants are enriched in AD, offering an independent line of evidence for biological relevance [18,19]. Integrating proteomic associations with orthogonal human genetic evidence therefore increases confidence that prioritized proteins are not merely correlates of disease state but candidate drivers or modifiers of disease onset [15,16,18,19].

Here, we applied a protected *APOE* ε4-focused framework to large-scale plasma proteomics. We enriched for older ε4 carriers who remained cognitively normal at ages when dementia risk is high in ε4 carriers, identified plasma proteins associated with this protected phenotype, and prioritized proteins supported by independent human genetic evidence. We hypothesized that proteins supported across proteomic and genetic analyses would highlight biological processes that actively delay clinical onset of AD.

## Methods

### Study design overview

We applied a protected-ε4-first plasma proteomic framework in the Global Neurodegeneration Proteomics Consortium (GNPC) to identify proteins associated with delayed clinical onset among *APOE*-ε4 carriers (**Figure 1**). The primary GNPC analysis compared older cognitively unimpaired ε4 carriers (“protected” ε4 carriers) with ε4 carriers affected by Alzheimer’s disease (AD). To contextualize ε4-specific findings within broader AD-related proteomic biology, we also performed population-wide plasma proteomic analyses across all *APOE* genotypes, modeling diagnostic categories together with *APOE* ε2 and ε4 dosage and *APOE* ε4-by-diagnosis interaction terms. Candidate proteins from the protected-ε4 analysis were subsequently integrated with external human genetic evidence from gene-based loss-of-function burden testing and Mendelian randomization.

**Figure 1.**
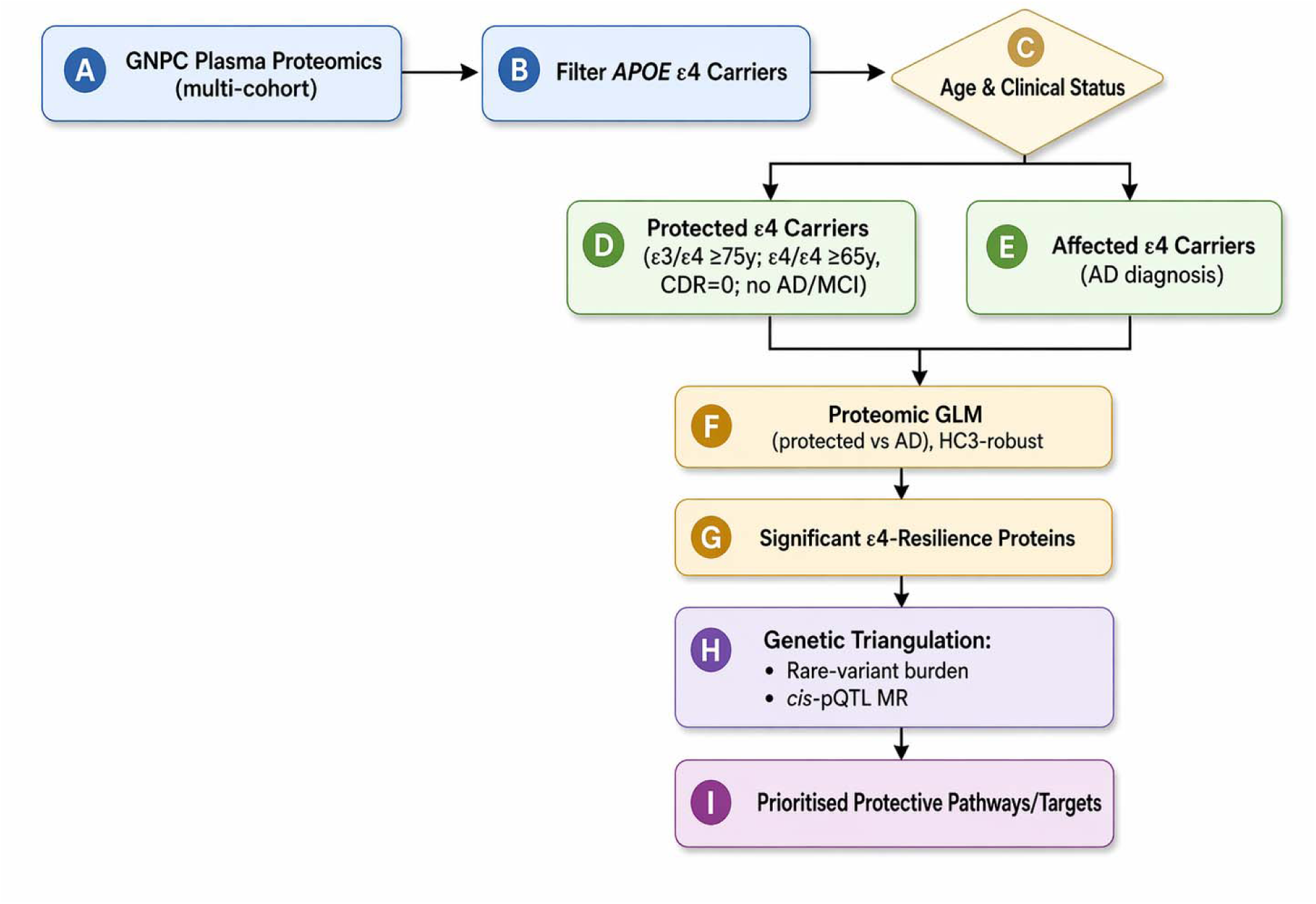
**Protected-**ε**4-first plasma proteomics and genetic triangulation framework.** Harmonized GNPC EDTA plasma proteomic data were used to compare clinically protected older *APOE*-ε4 carriers with *APOE*-ε4 carriers with Alzheimer’s disease. Protein-wise linear models identified protected-ε4-associated proteins, which were then interpreted in the context of population-wide diagnosis and *APOE* ε4-by-diagnosis analyses and prioritized through orthogonal human genetic evidence, including high-confidence loss-of-function burden testing and plasma/cerebrospinal fluid Mendelian randomization. GNPC, Global Neurodegeneration Proteomics Consortium.

### GNPC plasma proteomic cohorts and preprocessing

We used harmonized plasma proteomic data from multiple contributing GNPC cohorts. Analyses were restricted to EDTA plasma study samples. Protein measurements generated on SomaLogic SomaScan platforms were assembled into a unified plasma proteomic matrix, with removal of duplicate analyte columns and recoding-1 to missing. Based on the number of available analytes, samples were classified according to the SomaScan panel version (v3, v4, or v4.1).

### Clinical variable harmonization and diagnostic definitions

To improve comparability across contributing GNPC cohorts, we harmonized measures of clinical severity using Clinical Dementia Rating (CDR) whenever available. For participants without a recorded CDR value, we derived an approximate CDR category from available cognitive screening data using prespecified mappings for the Mini-Mental State Examination (MMSE) and Montreal Cognitive Assessment (MoCA). MMSE scores of 27–30, 21–26, 11–20, and 0–10 were mapped to CDR 0, 0.5, 1, and 2, respectively; the corresponding MoCA ranges were 26–30, 18–25, 11–17, and 0–10. Values coded as-1 were treated as missing.

To harmonize diagnoses across contributing cohorts, we used a rule-based approach centered on clinical severity while retaining a distinct category for non-AD neurodegenerative disorders. Participants with Parkinson’s disease, frontotemporal dementia, or amyotrophic lateral sclerosis were classified as non-AD neurodegenerative disease. For all other participants, final diagnostic assignment was based primarily on Clinical Dementia Rating (CDR): CDR = 0 defined cognitively normal controls, CDR = 0.5 defined mild cognitive impairment / subjective cognitive impairment (MCI/SCI), and CDR > 0.5 defined AD. If CDR was unavailable, we used the original cohort diagnosis variables, assigning AD when an AD diagnosis flag was present, MCI/SCI when an MCI/SCI flag was present, and control only when no disease flag was present and the available clinical information was consistent with normal cognition. This approach reduced between-cohort diagnostic heterogeneity, prioritized clinically interpretable severity categories, and preserved non-AD neurodegenerative disorders for secondary analyses.

### *APOE* harmonization and analytic cohorts

*APOE* genotype was harmonized across GNPC cohorts and represented as ε2 and ε4 allele dosages for regression analyses. Sex was similarly harmonized for use as a covariate. The primary protected-ε4 analysis focused on *APOE* ε3/ε4 and ε4/ε4 carriers. Protected ε4 carriers were defined as cognitively unimpaired participants at ages when dementia risk is expected to be high among ε4 carriers, specifically ε3/ε4 individuals aged 75 years or older and ε4/ε4 individuals aged 65 years or older. Affected ε4 carriers were defined using the same genotypes but included participants classified as having AD. Accordingly, the primary analysis compared protected versus affected ε4 carriers, capturing delayed clinical onset across the AD spectrum rather than AD alone. For secondary population-wide analyses, we included all participants with available plasma proteomics, complete covariate data, and one of four harmonized diagnostic categories: cognitively normal control, AD, MCI/SCI, or non-AD neurodegenerative disease. When repeated visits were available, the last available visit was retained for analysis.

### Plasma protein quality control, imputation, and principal components

Protein-level preprocessing was performed using the cleaned GNPC plasma proteomic matrix. Quality control followed an interquartile range (IQR)-based outlier detection strategy, in line with the GNPC plasma proteomics pipeline [20]. After outlier masking, samples and analytes were filtered according to call-rate thresholds, and remaining missing values were imputed using SoftImpute. Protein abundances were then transformed as log10(x+1). Principal component analysis was applied to the cleaned log-transformed matrix, with removal of principal component outliers followed by recomputation of the final proteomic principal components used as covariates in downstream models. All primary analyses reported here were based on the final IQR-QC, SoftImpute-completed, and PC-filtered plasma dataset.

### GNPC proteomic association analyses

For the primary protected-ε4 analysis, we performed protein-wise linear regression using HC3 robust standard errors, with log10-transformed plasma protein abundance as the dependent variable. The primary comparison was between protected and affected ε4 carriers, where affected carriers included participants with AD. Models were adjusted for ε4 allele dosage, age at visit, sex, and the first three plasma proteomic principal components. Multiple testing was controlled using the Benjamini–Hochberg false discovery rate (FDR).

For secondary population-wide analyses, we performed protein-wise linear regression across all *APOE* genotypes, again using log10-transformed plasma protein abundance as the outcome. Covariates included ε4 dosage, ε2 dosage, age at visit, sex, and harmonized diagnostic categories for AD, MCI/SCI, and non-AD neurodegenerative disease, with cognitively normal controls as the reference group. To assess whether proteomic associations differed according to ε4 status, we additionally included ε4-by-AD and ε4-by-MCI/SCI interaction terms. The first three plasma proteomic principal components were included in all models. These analyses provided the broader diagnosis-related and ε4-modified proteomic signals used for subsequent integration with the protected-ε4 analysis.

### Integration of protected-**ε**4 and population-wide proteomic signals

To identify the most robust candidate proteins, we integrated the protected-ε4 analysis with the broader GNPC population-wide proteomic analyses. Specifically, we considered three complementary domains of evidence: proteomic signals associated with AD-spectrum diagnosis, proteomic signals modified by ε4 status, and proteomic signals associated with protection among ε4 carriers. Within each analysis, statistical significance was defined using an FDR threshold of 0.05. We then evaluated concordance across analyses by examining whether proteins showed nominally significant associations in the same direction of effect across these complementary domains. This integration strategy was designed to prioritize proteins supported both by protected-ε4-specific and more general disease-related proteomic patterns.

### Loss-of-function burden analyses

To provide orthogonal human genetic support, we incorporated gene-based high-confidence loss-of-function burden results from a population-scale whole-genome sequencing analysis across UK Biobank, All of Us, and ADSP [19]. In that study, Alzheimer’s disease and dementia cases and controls were defined within each cohort using prespecified cohort-specific criteria [19], rare coding variants were annotated on GRCh38 using Ensembl VEP with LOFTEE [18] high-confidence loss-of-function classification, and gene-based burden testing was performed with REGENIE [21] with adjustment for age, sex, ancestry principal components, relatedness, and *APOE* ε4 and ε2 dosages. In the present manuscript, these results were used only as external gene-level support for prioritized GNPC proteins rather than as a primary discovery analysis.

### Mendelian randomization analyses

To provide orthogonal genetic support for prioritized proteins, we performed two-sample Mendelian randomization (MR) using protein quantitative trait loci (pQTLs) as exposure instruments and Alzheimer’s disease GWAS summary statistics [MVP preprint to be posted] as the outcome. Plasma pQTL instruments were derived from large-scale proteogenomic resources including deCODE SomaScan and UK Biobank Olink datasets [16], whereas cerebrospinal fluid (CSF) pQTL instruments were obtained from a large CSF proteogenomic meta-analysis [15].

For each protein, candidate pQTL instruments were selected from variants associated with protein abundance and present in the Alzheimer’s disease GWAS dataset. In the plasma analyses, instruments were required to meet a significance threshold of P < 5 × 10; in the CSF analyses, a more permissive threshold of P < 5 × 10 was used because of the smaller sample size of the CSF pQTL studies. To reduce redundancy and limit bias from correlated signals, instruments were restricted to one variant per 1 Mb region.

Exposure and outcome datasets were harmonized by effect allele before MR analysis. We focused primarily on inverse-variance weighted (IVW) estimates after exclusion of highly pleiotropic instruments, where pleiotropy was operationally defined as instrument variants associated with more than three proteins across the instrument-selection framework. When multiple instruments were available, MR results were generated using IVW together with sensitivity methods including weighted median and MR-Egger. When only a single instrument was available, the causal estimate was obtained using the Wald ratio.

For the current study, we prioritized IVW and Wald ratio results derived from non-pleiotropic instruments. These MR findings were then integrated with the GNPC proteomic and loss-of-function burden results to help prioritize proteins with convergent evidence for a potential role in delaying Alzheimer’s disease onset.

## Ethics and approvals

Each contributing cohort obtained approval from its local Institutional Review Board or Ethics Committee, and all participants gave written informed consent. Human studies were conducted in accordance with the Declaration of Helsinki. The current study protocol was granted an exemption by the Stanford University institutional review board because the analyses were carried out on deidentified data; therefore, additional informed consent was not required.

## Results

### Study population and analytic cohorts

After preprocessing and retention of participants present in the final IQR-based analytic datasets, the population-wide analysis included 9,734 individuals, comprising 4,659 cognitively unimpaired participants, 1,968 with MCI/SCI, 2,251 with AD, and 856 with non-AD neurodegenerative disease (**Supplementary Table S1**). In the protected-ε4 analysis, the current final uploaded analytic set included 456 protected ε4 carriers and 1,096 affected ε4 carriers with AD (**Table 1**). Across the broader GNPC sample, ε3/ε3 was the most common genotype, whereas ε4/ε4 carriers were enriched among AD cases relative to cognitively unimpaired participants. In the protected-ε4, unaffected participants were older than affected ε4 carriers by design, while the sex distribution was similar between groups.

**Table 1.**
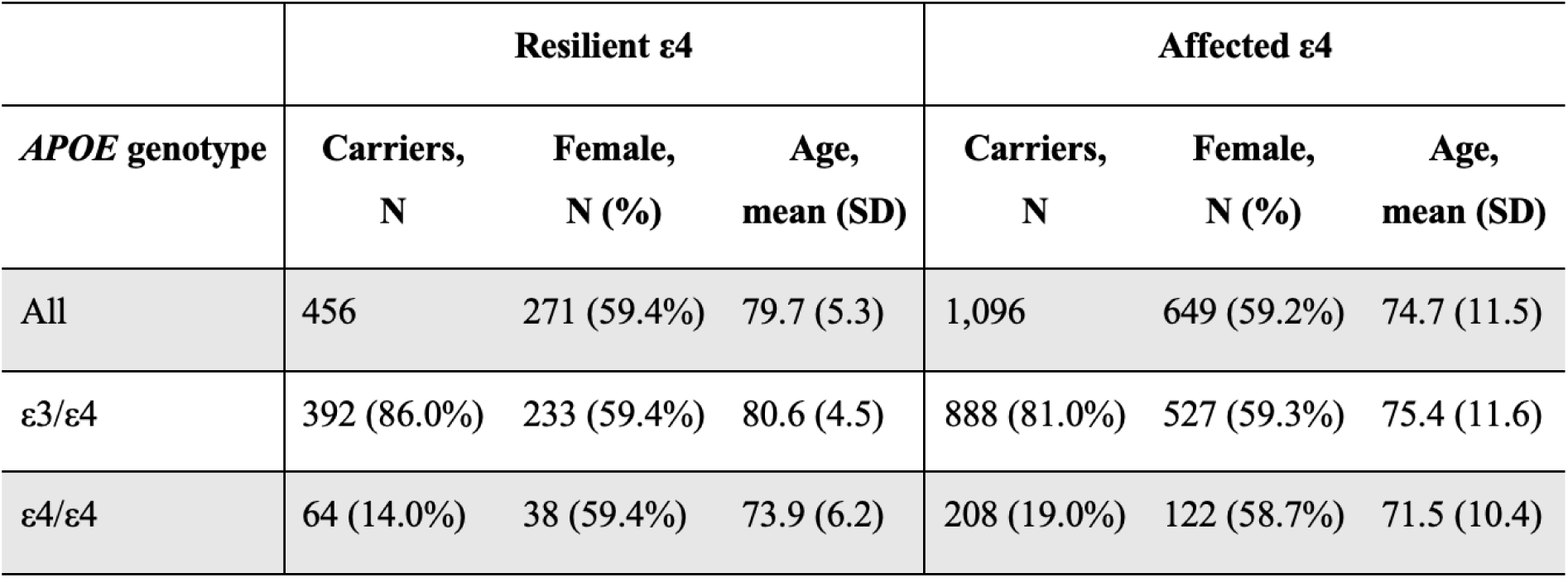
Demographics of the protected-ε4 analysis.

|  | **Resilient $\epsilon 4$** |  |  | **Affected $\epsilon 4$** |  |  |
| --- | --- | --- | --- | --- | --- | --- |
| ***APOE* genotype** | **Carriers,**<br>**N** | **Female,**<br>**N (%)** | **Age,**<br>**mean (SD)** | **Carriers,**<br>**N** | **Female,**<br>**N (%)** | **Age,**<br>**mean (SD)** |
| All | 456 | 271 (59.4%) | 79.7 (5.3) | 1,096 | 649 (59.2%) | 74.7 (11.5) |
| $\epsilon 3/\epsilon 4$ | 392 (86.0%) | 233 (59.4%) | 80.6 (4.5) | 888 (81.0%) | 527 (59.3%) | 75.4 (11.6) |
| $\epsilon 4/\epsilon 4$ | 64 (14.0%) | 38 (59.4%) | 73.9 (6.2) | 208 (19.0%) | 122 (58.7%) | 71.5 (10.4) |

### Proteome-wide landscape of protected-**ε**4-analysis

The protected-ε4 analysis identified a broad plasma proteomic signature associated with preserved cognition among older *APOE* ε4 carriers (**Figure 2; Supplementary Table S2**). In total, 721 of 7,588 assayed protein measures were associated with protected-ε4 at FDR < 0.05, including 386 proteins that were relatively higher in protected ε4 carriers and 335 that were relatively higher in affected ε4 carriers (**Supplementary Table S2**). Among the strongest signals enriched in protected ε4 carriers were NPTXR, SELENOW, ENO3, ENO2, LUM, NT5C, and NRCAM, whereas PLA2G7, SNAP23, TINAGL1, ODC1, AGRP, CD81, and APOB were among the strongest signals enriched in affected carriers.

**Figure 2.**
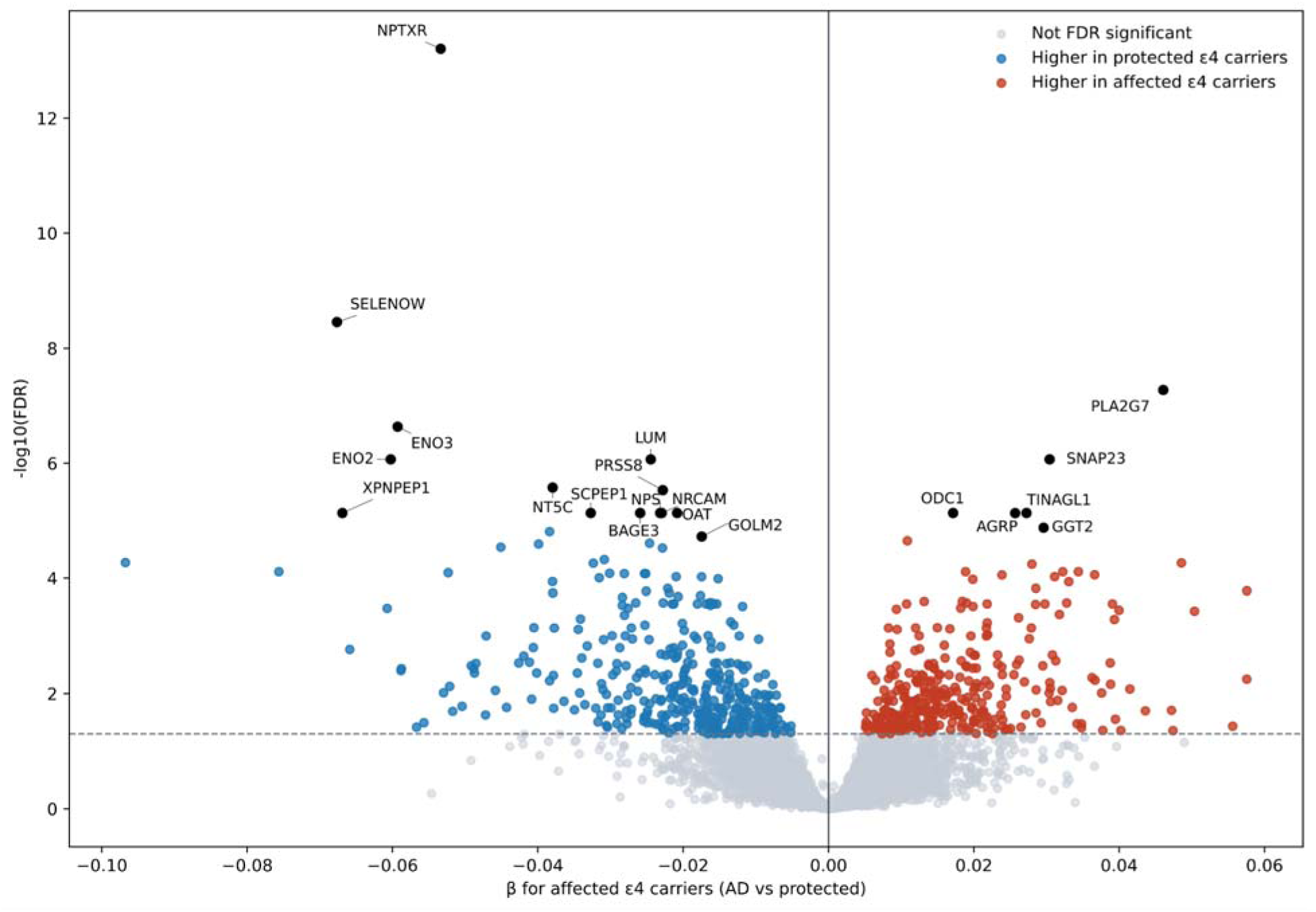
**Proteome-wide landscape of protected-**ε**4.** Volcano plot of the primary protected-ε4 analysis comparing protected *APOE* ε4 carriers with *APOE* ε4 carriers with Alzheimer’s disease. The x-axis shows the model effect estimate and the y-axis shows – log10(p). Negative β values indicate proteins relatively higher in protected ε4 carriers, whereas positive β values indicate proteins relatively higher in affected ε4 carriers. The horizontal dashed line denotes the false discovery rate threshold, and selected prioritized proteins are annotated.

Taken together, these results suggest that protected-ε4 is associated with widespread plasma protein differences rather than a limited set of isolated analytes. Broadly, proteins relatively higher in protected ε4 carriers included neuronal, synaptic, and structural candidates, whereas proteins relatively higher in affected ε4 carriers more often reflected inflammatory, lipid-related, and injury-associated biology.

### Integration with population-wide GNPC analyses

In the population-wide analysis, plasma proteomic differences associated with each of three clinical diagnoses versus controls were widespread (**Figure 3; Supplementary Tables S3 and S4**). AD was associated with 3,245 protein measures at FDR < 0.05, MCI/SCI with 3,153, and non-AD neurodegenerative disease with 3,857. In contrast, *APOE* ε4-by-diagnosis interaction effects were much more selective, with 130 significant protein measures for ε4-by-AD and only 13 for ε4-by-MCI/SCI (**Supplementary Tables S3 and S4**). These findings indicate that broad diagnosis-related proteomic differences are common in plasma, whereas *APOE* ε4-modified disease effects are comparatively restricted, supporting the value of the protected-ε4-first framework for enriching more specific biology.

**Figure 3.**
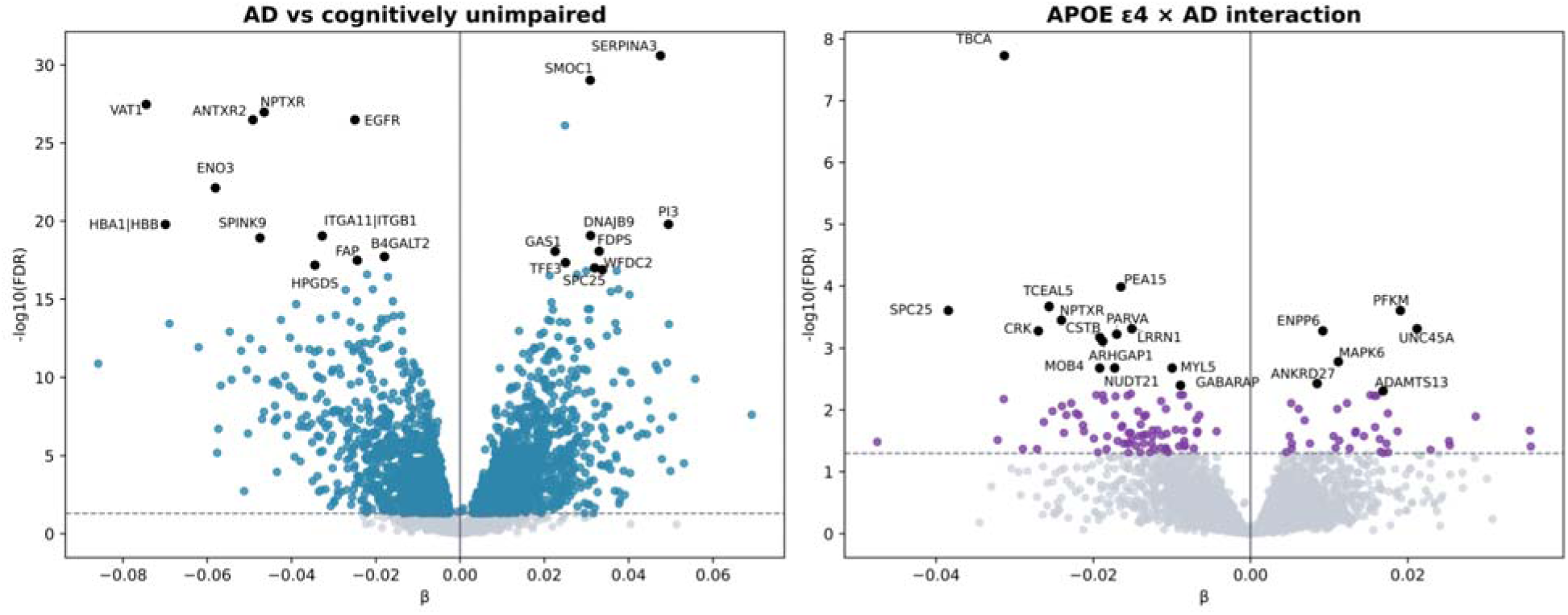
**Population-wide diagnosis and *APOE*-**ε**4 interaction analyses.** Left, volcano plot of the population-wide AD versus cognitively unimpaired comparison across the full GNPC sample. Right, volcano plot of the *APOE* ε4-by-AD interaction analysis. The x axis shows the model effect estimate and the y-axis shows –log10(p). Negative β values indicate lower protein levels in AD or in the *APOE* ε4-by-AD interaction term, and positive β values indicate higher levels. Selected prioritized proteins are annotated. AD, Alzheimer’s disease.

Among the strongest AD-associated proteins in the population-wide models, SERPINA3 and SMOC1 were increased in AD, whereas VAT1, NPTXR, ENO3, and ANTXR2 were decreased. The strongest ε4-by-AD interaction signals included TBCA, PEA15, TCEAL5, PFKM, SPC25, and NPTXR. Taken together, the population-wide analysis recapitulated broad AD-related plasma proteomic remodeling while also identifying a smaller subset of proteins whose disease associations differed according to *APOE*-ε4 status.

### Integration of protected-**ε**4 and population-wide signals

To place the protected-ε4 findings in the context of the broader GNPC disease-associated proteomic landscape, we examined overlap between proteins associated with protected-ε4 and those associated with AD or with ε4-by-AD interaction effects in the population-wide analysis. Of the 721 protein measures associated with protected-ε4 at FDR < 0.05, 458 were also significant in the population-wide AD model, and 440 of these showed concordant direction of effect **(Supplementary Table S5**). In addition, 49 were significant in the ε4-by-AD interaction model, and all 49 were directionally concordant with the protected-ε4 association. When false-discovery-rate correction was recalculated within the subset of protected-ε4-associated proteins, 474 remained significant for AD and 169 remained significant for the ε4-by-AD interaction, again with predominantly concordant directionality. Together, these findings indicate that a substantial proportion of the protected-ε4 signal overlaps with broader AD-related proteomic change, whereas a smaller subset appears to capture more selective ε4-modified disease biology (**Figure S1**).

This integrated analysis also helped distinguish proteins associated with general AD-related biology from those showing more specific evidence of ε4-modified disease association. For example, NPTXR, VAT1, ENO3, and SERPINA3 were identified both in the protected-ε4 analysis and in the broader AD case–control analysis, consistent with a relationship to general disease-associated proteomic change. By contrast, DBI, BPNT1, PCDH10, and one LILRA5 aptamer showed evidence of ε4-by-AD interaction, suggesting that their disease associations may be modified by ε4 status. These results provided the basis for subsequent prioritization of proteins supported across complementary lines of evidence.

### Orthogonal genetic support and candidate prioritization

Among proteins associated with protected-ε4, several showed convergent support across complementary analytic frameworks, including the population-wide GNPC models, gene-based high-confidence loss-of-function burden testing, and plasma or cerebrospinal fluid Mendelian randomization (**Figure 4**; **Table 2**). This convergence strengthens prioritization by identifying proteins supported not only by the protected-versus-affected ε4 comparison, but also by broader disease-related proteomic patterns and independent human genetic evidence.

**Figure 4.**
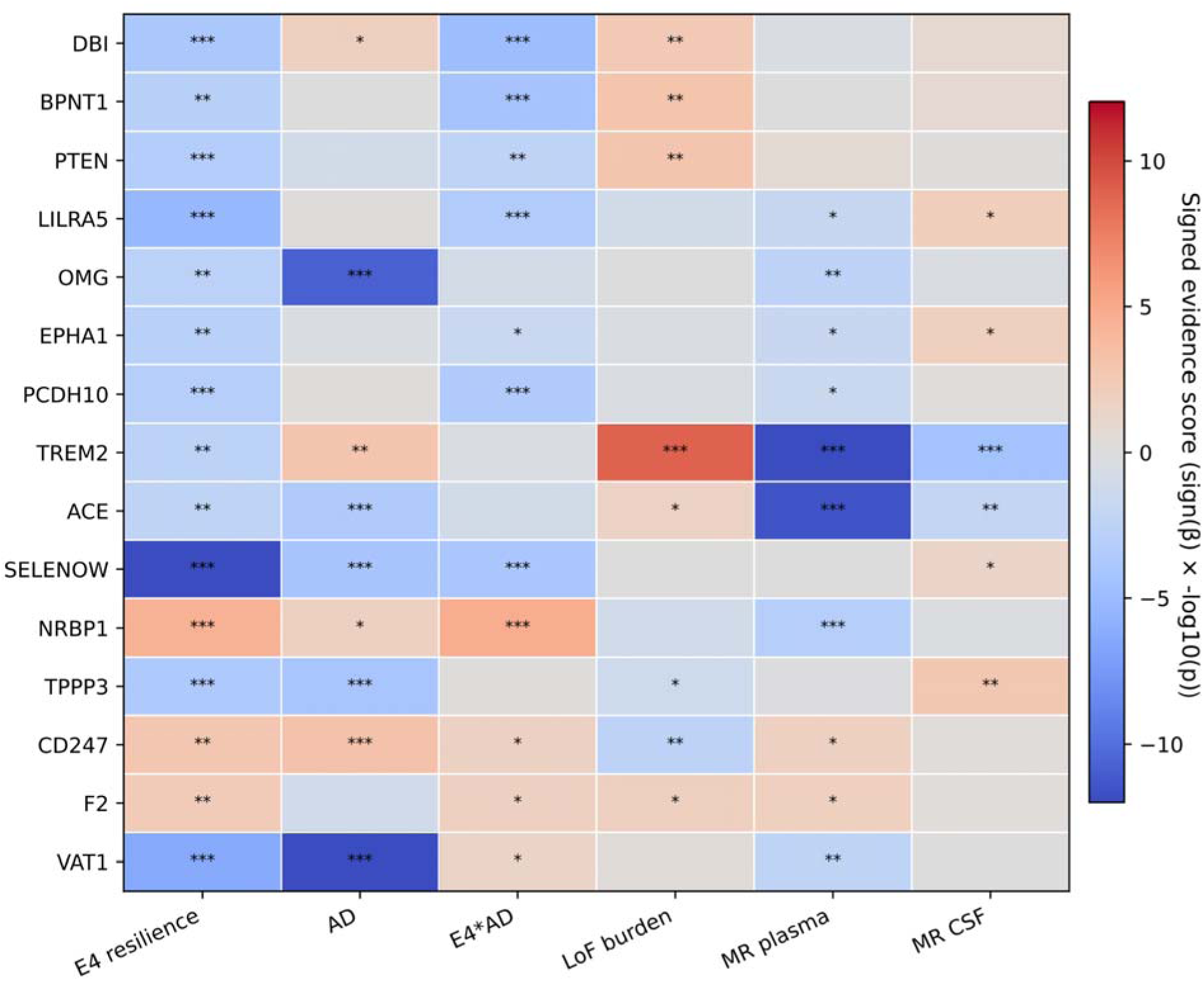
Integrated evidence across proteomic and genetic analyses. Heatmap summarizing direction and strength of evidence for prioritized proteins across the protected-ε4 analysis, the population-wide AD analysis, the *APOE* ε4-by-AD interaction analysis, high-confidence loss-of-function burden testing, and plasma/cerebrospinal fluid Mendelian randomization. Color indicates signed evidence score based on direction o effect and statistical strength. In-cell significance symbols denote nominal evidence thresholds: * P < 0.05, ** P < 0.01, and *** P < 0.001. For the proteomic analyses, false discovery rate significance is reported separately in the corresponding results tables and volcano plots. AD, Alzheimer’s disease; MR, Mendelian randomization.

**Table 2.** Detailed integrated summary of prioritized proteins across proteomic and genetic analyses. For each prioritized protein, the table reports the corresponding proteomic assay, effect estimates and p-values from the protected-ε4, AD, and ε4-by-AD analyses, high-confidence loss-of-function burden testing results, and the best available plasma and cerebrospinal fluid Mendelian randomization results.

| **Protein** | **ε4-Resilience** | **AD** | **ε4\*AD** | **LoF Burden** | **MR plasma** | **MR CSF** |
| --- | --- | --- | --- | --- | --- | --- |
| DBI | -0.020<br>(p=1.36e-04) | 0.008<br>(p=0.015) | -0.015<br>(p=1.52e-05) | 0.712<br>(p=0.004) | Wald: -0.016<br>(p=0.478) | IVW: 0.020<br>(p=0.154) |
| BPNT1 | -0.028<br>(p=0.001) | -0.000<br>(p=0.960) | -0.025<br>(p=5.93e-05) | 0.524<br>(p=0.001) | IVW: -0.006<br>(p=0.961) | IVW: 0.016<br>(p=0.220) |
| PTEN | -0.021<br>(p=5.56e-04) | -0.006<br>(p=0.103) | -0.012<br>(p=0.003) | 1.235<br>(p=0.002) | Wald: 0.163<br>(p=0.253) | IVW: 0.006<br>(p=0.733) |
| LILRA5 | -0.028<br>(p=5.20e-06) | 0.002<br>(p=0.666) | -0.017<br>(p=3.79e-04) | -0.258<br>(p=0.109) | IVW: -0.047<br>(p=0.013) | IVW: 0.031<br>(p=0.010) |
| OMG | -0.031<br>(p=0.002) | -0.040<br>(p=1.65e-11) | -0.010<br>(p=0.145) | 0.104<br>(p=0.812) | IVW: -0.213<br>(p=0.003) | IVW: -0.010<br>(p=0.362) |
| EPHA1 | -0.029<br>(p=0.002) | -0.004<br>(p=0.454) | -0.015<br>(p=0.018) | -0.045<br>(p=0.509) | IVW: -0.119<br>(p=0.017) | IVW: 0.019<br>(p=0.012) |
| PCDH10 | -0.023<br>(p=6.53e-04) | 0.002<br>(p=0.719) | -0.018<br>(p=2.83e-04) | -0.386<br>(p=0.407) | IVW: -0.017<br>(p=0.026) | IVW: 0.011<br>(p=0.584) |
| TREM2 | -0.025<br>(p=0.002) | 0.018<br>(p=0.001) | -0.006<br>(p=0.346) | 0.931<br>(p=1.27e-09) | IVW: -0.142<br>(p=3.75e-23) | IVW: -0.125<br>(p=4.86e-05) |
| ACE | -0.018<br>(p=0.003) | -0.014<br>(p=2.57e-04) | -0.007<br>(p=0.111) | 0.171<br>(p=0.031) | IVW: -0.145<br>(p=2.65e-12) | IVW: -0.080<br>(p=0.008) |
| SELENOW | -0.068<br>(p=9.23e-13) | -0.024<br>(p=6.24e-05) | -0.026<br>(p=1.13e-04) | 0.045<br>(p=0.833) |  | Wald: 0.086<br>(p=0.044) |
| NRBP1 | 0.020<br>(p=4.29e-05) | 0.007<br>(p=0.019) | 0.015<br>(p=1.80e-05) | -1.213<br>(p=0.077) | IVW: -0.088<br>(p=7.11e-04) | IVW: -0.010<br>(p=0.596) |
| TPPP3 | -0.019<br>(p=1.95e-04) | -0.013<br>(p=9.20e-05) | 0.001<br>(p=0.736) | -0.515<br>(p=0.041) | IVW: -0.028<br>(p=0.745) | IVW: 0.049<br>(p=0.002) |
| CD247 | 0.018<br>(p=0.002) | 0.012<br>(p=6.73e-04) | 0.009<br>(p=0.024) | -0.434<br>(p=0.003) | Wald: 0.251<br>(p=0.019) | IVW: 0.008<br>(p=0.539) |
| F2 | 0.008<br>(p=0.005) | -0.003<br>(p=0.085) | 0.005<br>(p=0.020) | 0.550<br>(p=0.015) | IVW: 0.083<br>(p=0.011) | IVW: 0.008<br>(p=0.605) |
| VAT1 | -0.052<br>(p=3.83e-07) | -0.074<br>(p=1.33e-31) | 0.015<br>(p=0.040) | 0.189<br>(p=0.441) | IVW: -0.097<br>(p=0.003) | IVW: 0.002<br>(p=0.887) |

A first group of proteins combined protected-ε4 association with evidence of ε4-modified disease effects in the population-wide models. These included DBI, BPNT1, PTEN, LILRA5, EPHA1, PCDH10, NRBP1, CD247, and F2, suggesting that their associations are not limited to general case–control differences but may be particularly relevant to ε4-associated disease biology. Within this set, DBI, BPNT1, and PTEN also showed supportive high-confidence loss-of-function burden results, whereas LILRA5, EPHA1, PCDH10, NRBP1, CD247, and F2 had additional support from plasma and/or CSF Mendelian randomization. These proteins were not uniform in direction: DBI, BPNT1, PTEN, LILRA5, EPHA1, and PCDH10 were relatively higher in protected ε4 carriers, whereas NRBP1, CD247, and F2 were relatively higher in affected ε4 carriers.

A second group aligned more closely with broader AD-related proteomic change. OMG, ACE, SELENOW, VAT1, and TPPP3 were all associated with protected-ε4 and also showed lower levels in AD in the population-wide analysis. Within this set, ACE, OMG, VAT1, and TPPP3 had additional support from plasma or CSF Mendelian randomization, and TPPP3 also showed supportive loss-of-function burden evidence. In addition, TREM2 provided a strong positive-control signal, showing association with protected-ε4 together with robust support from loss-of-function burden testing and both plasma and CSF Mendelian randomization.

Taken together, these findings suggest that the protected-ε4-first framework captures at least two complementary patterns (**Figure 4; Figure S2**): proteins that may reflect broader AD-related biology and proteins that may be more informative for ε4-specific disease modification. Within the latter group, LILRA5 is particularly notable because it combined protected-ε4 association, evidence of ε4-by-AD interaction in the GNPC models, and supportive Mendelian randomization. Other proteins with a similar ε4-modified pattern, including DBI, BPNT1, PTEN, EPHA1, and PCDH10, may therefore be especially promising as candidate modifiers of delayed onset among ε4 carriers.

## Discussion

The central contribution of this study is a protected-ε4-first design that focuses on biological mechanisms associated with preserved clinical status in older *APOE*-ε4 carriers, rather than on proteins that simply distinguish cases from controls [12]. This distinction matters in AD plasma proteomics because many case–control signals are likely to reflect downstream neurodegeneration, frailty, vascular comorbidity, or treatment exposure rather than mechanisms that actively delay symptom onset. The broader GNPC literature makes this framing biologically plausible, as carrying *APOE*-ε4 is associated with a conserved immune-related proteomic signature across neurodegenerative diseases, implying that ε4 creates a basal vulnerability state that still requires additional modifiers to shape clinical expression [12,22]. In this context, the main value of the present results lies in distinguishing proteins that primarily reflect broader AD-related biology from those that may be more informative for ε4-specific disease modification [22].

Within the ε4-modified tier, LILRA5 is the most immediately compelling hypothesis-generating candidate. In our data, LILRA5 combined protected-ε4 association, ε4-by-AD interaction, and supportive Mendelian randomization. The relevance of that pattern is strengthened by a recent large *APOE*-stratified GWAS, which identified a genome-wide significant signal at the *LILRA5* locus in the ε4-positive stratum [23]. Together, these findings highlight convergence between proteomic ε4-by-AD interaction and genotype-stratified human genetics at this locus. Biologically, *LILRA5* belongs to the leukocyte immunoglobulin-like receptor cluster on chromosome 19, and recent work shows that *LILRA5* is expressed in naïve monocytes and neutrophils and can trigger reactive oxygen species generation and inflammatory signaling [24,25]. In a disease context in which *APOE*-ε4 already appears to prime chronic immune dysregulation, *LILRA5* is therefore a plausible candidate for ε4-conditioned innate immune amplification rather than merely a generic inflammatory marker [22].

*EPHA1* belongs close to *LILRA5* in this interpretive tier, although the underlying biology is different. *EPHA1* is an established AD susceptibility locus from large GWAS, and functional follow-up of the AD-associated P460L variant suggests altered receptor activity with consequences for endothelial behavior and blood–brain barrier function [26]. That is relevant here because BBB dysfunction is increasingly recognized as part of AD pathophysiology, and a protected-ε4-associated EPHA1 signal could plausibly index neurovascular protection rather than only amyloid-or tau-centered biology [27]. *PTEN* is also compelling because its biology sits at the intersection of synaptic signaling, neuronal energetics, and amyloid-related synaptic toxicity [28]. Experimental work has shown that Aβ can recruit PTEN to postsynaptic compartments and drive synaptic depression, while human tissue data suggest that aberrant synaptic PTEN increases with symptomatic AD progression [28]. In the present setting, *PTEN* therefore fits a model in which preserved signaling homeostasis and resistance to synaptic depression may help postpone threshold crossing in ε4 carriers.

PCDH10 adds a complementary synaptic and circuit-maintenance dimension. *PCDH10* is a protocadherin involved in excitatory synapse development and circuit refinement [29], its association here is consistent with the possibility that protected-ε4 resilience may involve preservation of synaptic architecture in addition to modulation of inflammatory pathways. DBI and BPNT1 are less established in AD, but both are mechanistically plausible and therefore remain biologically relevant candidates in the present framework. *DBI* encodes diazepam-binding inhibitor/acyl-CoA-binding protein, a neuropeptide with links to GABAergic signaling and lipid metabolism, and older clinical work reported altered CSF DBI levels in AD and other dementias. BPNT1 is a lithium-sensitive 3′-phosphoadenosine 5′-phosphate phosphatase with roles in sulfation-related nucleotide metabolism [30]. Experimental work has linked BPNT1 inhibition to neuronal dysfunction and lithium-responsive biology [31]. *DBI* and *BPNT1* are not established AD susceptibility genes, but their known biology supports their candidacy as markers of metabolic-stress and neuromodulatory pathways that may be underappreciated in current AD target prioritization.

A second set of proteins appears to align more closely with broader AD-related plasma remodeling, but several of them remain highly informative. OMG, reduced here in AD versus controls. is particularly important because a new multi-cohort study now supports it as a brain-specific proteomic determinant of neurodegenerative resiliency [32]. In that study, lower plasma OMG was associated with cortical amyloid deposition, compromised brain structure, dementia, and future dementia risk, and genetic analyses supported OMG as causally protective across several neurodegenerative outcomes [32]. This independent evidence strengthens the interpretation of the present OMG finding and supports its relevance to protected-ε4 resilience or resistance-related biology. SELENOW is also noteworthy because recent experimental work showed that selenoprotein W modulates tau homeostasis and synaptic maintenance in an AD mouse model [33]. VAT1 is less AD-specific, but it is a neuronal vesicle protein with oxidoreductase activity and links to vesicular transport, mitochondrial fusion, and phospholipid biology[34], making lower VAT1 in AD and relative preservation in protected ε4 carriers biologically plausible. TPPP3 remains a limited-evidence but intriguing candidate, as it belongs to a tubulin polymerization-promoting family and has been linked to axon regeneration [35] and microtubule biology [36], which is relevant to neurodegeneration even if direct AD literature is still sparse. Together, OMG, SELENOW, VAT1, and TPPP3 support the possibility that protected-ε4 resilience or resistance is partly mediated by preservation of axonal integrity, synaptic structure, and cytoskeletal stability [32–35].

Three proteins merit a more nuanced interpretation because they were relatively higher in affected rather than protected ε4 carriers: NRBP1, CD247, and F2. In our data, all three also showed interaction and/or orthogonal genetic support, arguing against a simplistic “lower in affected equals risk-increasing” model. NRBP1 is mechanistically relevant because it regulates degradation of BRI2 and BRI3, physiological inhibitors of APP processing and Aβ oligomerization, and NRBP1 depletion reduces Aβ production in neuronal cells [37]. CD247 is the T-cell receptor ζ-chain, and although it is not an established AD risk gene, its appearance fits a growing literature implicating adaptive immune [38] and CD8+ T-cell alterations in AD progression [39]. F2 encodes prothrombin, and thrombin is increasingly viewed as a neurovascular and inflammatory mediator in AD that can contribute to BBB dysfunction, microglial activation, and neurotoxicity [40–42]. These genes may therefore prove most useful as markers of ε4-conditioned vulnerability pathways, compensatory responses, or context-dependent amplifiers rather than straightforward protective mediators.

TREM2 and ACE occupy a different interpretive category from the less established candidates. In particular, *TREM2* is already strongly anchored in AD pathobiology: rare coding variants in *TREM2* were among the earliest non-*APOE* genetic associations and to implicate microglial lipid handling and phagocytic function in AD [43]. Subsequent CSF proteogenomic studies have reinforced the relevance of *TREM2*-related biology to disease mechanisms [44]. ACE plays a similar positive-control role it is a late-onset AD GWAS locus [45] and can convert Aβ42 to Aβ40 [46,47], with *ACE* inhibition enhancing amyloid deposition in experimental systems. The re-emergence of TREM2 and ACE is therefore reassuring, because it shows that the protected-ε4-first framework recovers established AD-relevant biology.

From a translational perspective, the most promising proteins are not necessarily those with the largest effect sizes, but those with the clearest convergence across protected-ε4 association, ε4-modified disease association, and human genetic support. On current evidence, LILRA5 stands out as the clearest ε4-specific hypothesis-generating candidate, with EPHA1, PTEN, DBI, BPNT1, and PCDH10 forming a second tier of plausible modifiers, while OMG, SELENOW, VAT1, and TPPP3 appear especially informative for a broader axonal and structural-resilience axis. The next steps should be replication in independent ε4-enriched cohorts, longitudinal validation that these proteins track delayed conversion rather than survivor bias, and integration with amyloid and tau biomarkers to distinguish resistance-like from resilience-like mechanisms of protection. More broadly, these data support a meaningful shift towards prevention-focused AD target discovery.

## Data Availability

GNPC data available on the ADworkbench.
Genomics related summary statistics are available through their referenced corresponding publications.

https://addi-portal.alzheimersdata.org

## Acknowledgments

This work was supported by the National Institute of Health and National Institute of Aging grants AG072290 (MDG), AG066515 (MDG).

We thank the participants, families, investigators, staff, and contributing cohorts of the Global Neurodegeneration Proteomics Consortium (GNPC), whose biospecimens, clinical data, and proteomic data made this work possible. The GNPC harmonized dataset was accessed and analyzed through the Alzheimer’s Disease Data Initiative AD Workbench. Data discovery and/or analysis services contributing to this work were provided in-kind by the AD Data Initiative. We also acknowledge Gates Ventures and Johnson & Johnson for supporting the majority of GNPC biosample proteomic analyses, as described in the GNPC resource publication.

## Declaration of Competing Interest

The authors declare that they have no known competing financial interests or personal relationships that could have appeared to influence the work reported in this manuscript.

## Supplementary tables

**Table S1. Demographic characteristics of the overall GNPC analytic cohort.** The table summarizes age, sex, and *APOE* genotype distribution across cognitively unimpaired, MCI/SCI, AD, and non-AD neurodegenerative disease groups in the population-wide GNPC analysis. Values are presented as number, number (percentage), or mean (standard deviation), as appropriate.

**Table S2. FDR-significant proteins in the *APOE*** ε**4 protected analysis.** Full list of protein measures associated with the protected-versus-affected *APOE* ε4 comparison at false discovery rate <0.05, including effect estimates, standard errors, p values, and adjusted p values.

**Table S3. FDR-significant proteins in the population-wide AD analysis.** Full list of protein measures associated with AD versus cognitively unimpaired status at false discovery rate <0.05 in the population-wide GNPC analysis, including effect estimates, standard errors, p values, and adjusted p values.

**Table S4. FDR-significant proteins in the population-wide *APOE*** ε**4-by-AD interaction analysis.** Full list of protein measures showing significant *APOE* ε4-by-AD interaction effects at false discovery rate <0.05, including effect estimates, standard errors, p values, and adjusted p values.

**Table S5. Overlap of *APOE*** ε**4 protected, AD, and *APOE*** ε**4-by-AD signals.** Protein-level overlap between the *APOE* ε4 protected analysis, the population-wide AD analysis, and the *APOE* ε4-by-AD interaction analysis, including direction-of-effect concordance across analyses.

---

**Figure S1.**
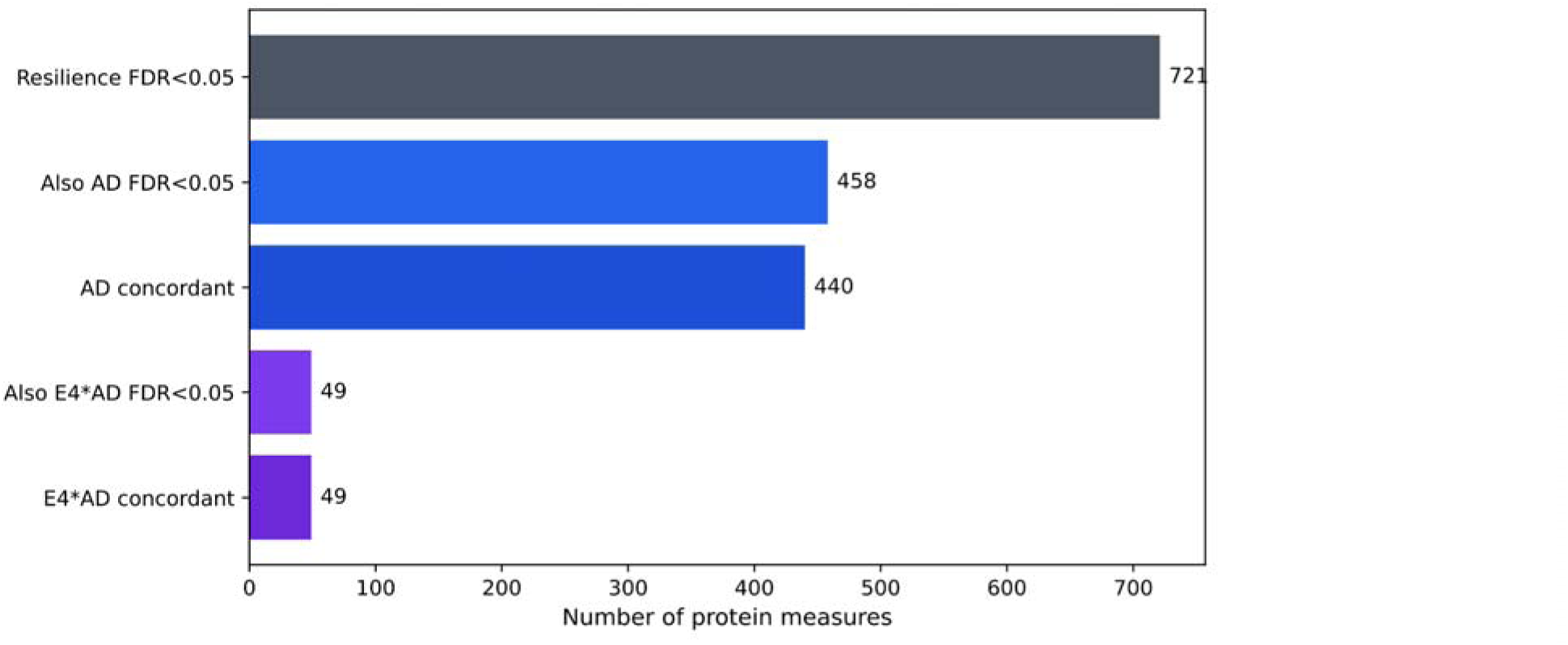
**Overlap of *APOE*-**ε**4 protected signals with population-wide AD and *APOE*** ε**4-by-AD signals.** Summary of the overlap between proteins associated with *APOE*-ε4 protected and proteins associated with broader AD-related proteomic change or *APOE* ε4-modified disease effects in the population-wide analysis. This figure illustrates the extent to which protected-associated proteins map onto general AD biology versus more selective ε4-modified signals.

**Figure S2.**
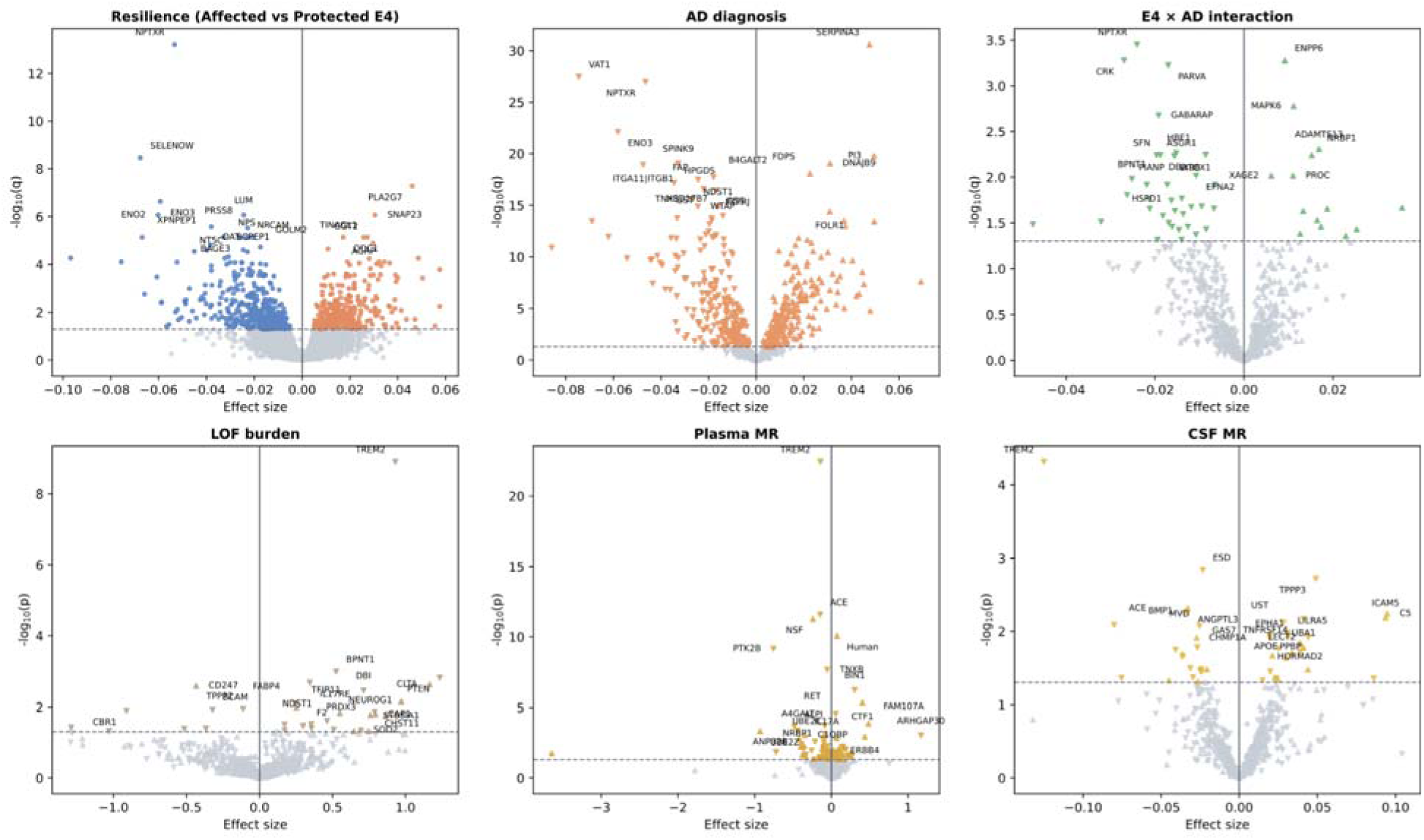
**Integrated view of *APOE*-**ε**4 protected associated proteins across case–control, interaction, and genetic analyses.** Six volcano plots summarize evidence across the primary *APOE*-ε4 protected analysis, the population-wide AD diagnosis analysis, the *APOE* ε4-by-AD interaction analysis, high-confidence loss-of-function burden testing, and plasma and cerebrospinal fluid Mendelian randomization. In the protected panel, all tested proteins are shown, with proteins relatively higher in protected ε4 carriers shown as downward triangles and proteins relatively higher in affected ε4 carrier shown as upward triangles. In the remaining panels, plotting is restricted to proteins that were significant in the *APOE*-ε4 protected analysis at false discovery rate <0.05, and the triangle direction is carried forward from the protected analysis to facilitate visual comparison across analytic frameworks. Colored symbols denote nominally significant associations within each panel, whereas gray symbols denote non-significant associations. The x-axis shows the signed effect estimate and the y-axis shows –log10(q) for proteomic analyses or –log10(p) for burden and Mendelian randomization analyses. Selected top-ranking proteins are annotated in each panel. AD, Alzheimer’s disease; CSF, cerebrospinal fluid; MR, Mendelian randomization.

## Notes

### Competing Interest Statement

The authors have declared no competing interest.

### Author Declarations

Each contributing cohort obtained approval from its local Institutional Review Board or Ethics Committee, and all participants gave written informed consent. Human studies were conducted in accordance with the Declaration of Helsinki. The current study protocol was granted an exemption by the Stanford University institutional review board because the analyses were carried out on deidentified data; therefore, additional informed consent was not required. https://www.neuroproteome.org/harmonized-data-set-hds

## References

[1] 2025 Alzheimer’s disease facts and figures. Alzheimers Dement 2025;21:e70235. 10.1002/alz.70235.

[2] Gustavsson A, Norton N, Fast T, Frölich L, Georges J, Holzapfel D, et al. Global estimates on the number of persons across the Alzheimer’s disease continuum. Alzheimers Dement 2023;19:658–70. 10.1002/alz.12694.

[3] Long JM, Holtzman DM. Alzheimer Disease: An Update on Pathobiology and Treatment Strategies. Cell 2019;179:312–39. 10.1016/j.cell.2019.09.001.

[4] Jack CR, Bennett DA, Blennow K, Carrillo MC, Dunn B, Haeberlein SB, et al. NIA-AA Research Framework: Toward a biological definition of Alzheimer’s disease. Alzheimer’s & Dementia 2018;14:535–62. 10.1016/j.jalz.2018.02.018.

[5] Belloy ME, Andrews SJ, Le Guen Y, Cuccaro M, Farrer LA, Napolioni V, et al. APOE Genotype and Alzheimer Disease Risk Across Age, Sex, and Population Ancestry. JAMA Neurology 2023;80:1284–94. 10.1001/jamaneurol.2023.3599.

[6] Fortea J, Pegueroles J, Alcolea D, Belbin O, Dols-Icardo O, Vaqué-Alcázar L, et al. APOE4 homozygosity represents a distinct genetic form of Alzheimer’s disease. Nat Med 2024;30:1284–91. 10.1038/s41591-024-02931-w.

[7] Arenaza-Urquijo EM, Vemuri P. Resistance vs resilience to Alzheimer disease. Neurology 2018;90:695–703. 10.1212/WNL.0000000000005303.

[8] Kaup AR, Nettiksimmons J, Harris TB, Sink KM, Satterfield S, Metti AL, et al. Cognitive Resilience to Apolipoprotein E ε4: Contributing Factors in Black and White Older Adults. JAMA Neurol 2015;72:340–8. 10.1001/jamaneurol.2014.3978.

[9] Pappalettera C, Carrarini C, Cappa S, Caraglia N, Cotelli M, Marra C, et al. Challenges to identifying risk versus protective factors in Alzheimer’s disease. Nat Med 2024;30:3094–5. 10.1038/s41591-024-03158-5.

[10] Gold L, Ayers D, Bertino J, Bock C, Bock A, Brody EN, et al. Aptamer-based multiplexed proteomic technology for biomarker discovery. PLoS One 2010;5:e15004. 10.1371/journal.pone.0015004.

[11] Kim CH, Tworoger SS, Stampfer MJ, Dillon ST, Gu X, Sawyer SJ, et al. Stability and reproducibility of proteomic profiles measured with an aptamer-based platform. Sci Rep 2018;8:8382. 10.1038/s41598-018-26640-w.

[12] Imam F, Saloner R, Vogel JW, Krish V, Abdel-Azim G, Ali M, et al. The Global Neurodegeneration Proteomics Consortium: biomarker and drug target discovery for common neurodegenerative diseases and aging. Nat Med 2025;31:2556–66. 10.1038/s41591-025-03834-0.

[13] Guo Y, You J, Zhang Y, Liu W-S, Huang Y-Y, Zhang Y-R, et al. Plasma proteomic profiles predict future dementia in healthy adults. Nat Aging 2024;4:247–60. 10.1038/s43587-023-00565-0.

[14] Emilsson V, Ilkov M, Lamb JR, Finkel N, Gudmundsson EF, Pitts R, et al. Co-regulatory networks of human serum proteins link genetics to disease. Science 2018;361:769–73. 10.1126/science.aaq1327.

[15] Western D, Timsina J, Wang L, Wang C, Yang C, Phillips B, et al. Proteogenomic analysis of human cerebrospinal fluid identifies neurologically relevant regulation and implicates causal proteins for Alzheimer’s disease. Nat Genet 2024;56:2672–84. 10.1038/s41588-024-01972-8.

[16] Eldjarn GH, Ferkingstad E, Lund SH, Helgason H, Magnusson OT, Gunnarsdottir K, et al. Large-scale plasma proteomics comparisons through genetics and disease associations. Nature 2023;622:348–58. 10.1038/s41586-023-06563-x.

[17] Hemani G, Zheng J, Elsworth B, Wade KH, Haberland V, Baird D, et al. The MR-Base platform supports systematic causal inference across the human phenome. Elife 2018;7:e34408. 10.7554/eLife.34408.

[18] Karczewski KJ, Francioli LC, Tiao G, Cummings BB, Alföldi J, Wang Q, et al. The mutational constraint spectrum quantified from variation in 141,456 humans. Nature 2020;581:434–43. 10.1038/s41586-020-2308-7.

[19] Le Guen Y, Peña-Tauber A, Pulgrossi RC, Park J, Orias H, Greicius MD. Population-scale burden analysis of rare damaging coding variants identifies novel risk genes for Alzheimer’s disease and Parkinson’s disease 2026:2026.03.03.26347540. 10.64898/2026.03.03.26347540.

[20] Ali M, Erabadda B, Chen Y, Xu Y, Gong K, Liu M, et al. Shared and disease-specific pathways in frontotemporal dementia and Alzheimer’s and Parkinson’s diseases. Nat Med 2025;31:2567–77. 10.1038/s41591-025-03833-1.

[21] Mbatchou J, Barnard L, Backman J, Marcketta A, Kosmicki JA, Ziyatdinov A, et al. Computationally efficient whole-genome regression for quantitative and binary traits. Nat Genet 2021;53:1097–103. 10.1038/s41588-021-00870-7.

[22] Shvetcov A, Johnson ECB, Winchester LM, Walker KA, Wilkins HM, Thompson TG, et al. APOE ε4 carriers share immune-related proteomic changes across neurodegenerative diseases. Nat Med 2025;31:2590–601. 10.1038/s41591-025-03835-z.

[23] Thomassen JQ, Leonard H, Ulms B, Grenier-Boley B, Heikkinen S, Garcia-González P, et al. APOE stratified genome-wide association studies provide novel insights into the genetic etiology of Alzheimers’s disease 2025:2025.05.07.25327065. 10.1101/2025.05.07.25327065.

[24] Fu Z, Rumpret M, Kube-Golovin I, Lyndin M, Solntceva V, Zhao Y, et al. LILRA5 Functions to Induce ROS Production on Innate Immune Cells. Eur J Immunol 2025;55:e70079. 10.1002/eji.70079.

[25] Mitchell A, Rentero C, Endoh Y, Hsu K, Gaus K, Geczy C, et al. LILRA5 is expressed by synovial tissue macrophages in rheumatoid arthritis, selectively induces pro-inflammatory cytokines and IL-10 and is regulated by TNF-α, IL-10 and IFN-γ. European Journal of Immunology 2008;38:3459–73. 10.1002/eji.200838415.

[26] Owens HA, Thorburn LE, Walsby E, Moon OR, Rizkallah P, Sherwani S, et al. Alzheimer’s disease-associated P460L variant of EphA1 dysregulates receptor activity and blood-brain barrier function. Alzheimers Dement 2024;20:2016–33. 10.1002/alz.13603.

[27] Preis L, Villringer K, Brosseron F, Düzel E, Jessen F, Petzold GC, et al. Assessing blood-brain barrier dysfunction and its association with Alzheimer’s pathology, cognitive impairment and neuroinflammation. Alz Res Therapy 2024;16:172. 10.1186/s13195-024-01529-1.

[28] Knafo S, Sánchez-Puelles C, Palomer E, Delgado I, Draffin JE, Mingo J, et al. PTEN recruitment controls synaptic and cognitive function in Alzheimer’s models. Nat Neurosci 2016;19:443–53. 10.1038/nn.4225.

[29] Hoshina N, Johnson-Venkatesh EM, Rally VR, Sant J, Hoshina M, Seiglie MP, et al. ASD/OCD-Linked Protocadherin-10 Regulates Synapse, But Not Axon, Development in the Amygdala and Contributes to Fear-and Anxiety-Related Behaviors. J Neurosci 2022;42:4250–66. 10.1523/JNEUROSCI.1843-21.2022.

[30] Ferrarese C, Appollonio I, Frigo M, Meregalli S, Piolti R, Tamma F, et al. Cerebrospinal fluid levels of diazepam-binding inhibitor in neurodegenerative disorders with dementia. Neurology 1990;40:632–5. 10.1212/wnl.40.4.632.

[31] Meisel JD, Kim DH. Inhibition of Lithium-Sensitive Phosphatase BPNT-1 Causes Selective Neuronal Dysfunction in *C. elegans*. Current Biology 2016;26:1922–8. 10.1016/j.cub.2016.05.050.

[32] Duggan MR, Oh HS-H, Frank P, Gomez GT, Zweibaum D, Cui Y, et al. OMG! A proteomic determinant of neurodegenerative resiliency. Mol Neurodegeneration 2026;21:9. 10.1186/s13024-025-00921-1.

[33] Ren B, Situ J, Huang X, Tan Q, Xiao S, Li N, et al. Selenoprotein W modulates tau homeostasis in an Alzheimer’s disease mouse model. Commun Biol 2024;7:872. 10.1038/s42003-024-06572-0.

[34] Kim S-Y, Mori T, Chek MF, Furuya S, Matsumoto K, Yajima T, et al. Structural insights into vesicle amine transport-1 (VAT-1) as a member of the NADPH-dependent quinone oxidoreductase family. Sci Rep 2021;11:2120. 10.1038/s41598-021-81409-y.

[35] Rao M, Luo Z, Liu C-C, Chen C-Y, Wang S, Nahmou M, et al. Tppp3 is a novel molecule for retinal ganglion cell identification and optic nerve regeneration. Acta Neuropathol Commun 2024;12:204. 10.1186/s40478-024-01917-6.

[36] Oláh J, Lehotzky A, Szénási T, Berki T, Ovádi J. Modulatory Role of TPPP3 in Microtubule Organization and Its Impact on Alpha-Synuclein Pathology. Cells 2022;11:3025. 10.3390/cells11193025.

[37] Yasukawa T, Tsutsui A, Tomomori-Sato C, Sato S, Saraf A, Washburn MP, et al. NRBP1-Containing CRL2/CRL4A Regulates Amyloid β Production by Targeting BRI2 and BRI3 for Degradation. Cell Reports 2020;30:3478–3491.e6. 10.1016/j.celrep.2020.02.059.

[38] van Olst L, Kamermans A, Halters S, van der Pol SMA, Rodriguez E, Verberk IMW, et al. Adaptive immune changes associate with clinical progression of Alzheimer’s disease. Mol Neurodegeneration 2024;19:38. 10.1186/s13024-024-00726-8.

[39] Hu D, Weiner HL. Unraveling the dual nature of brain CD8+ T cells in Alzheimer’s disease. Mol Neurodegeneration 2024;19:16. 10.1186/s13024-024-00706-y.

[40] Festoff BW, Sajja RK, van Dreden P, Cucullo L. HMGB1 and thrombin mediate the blood-brain barrier dysfunction acting as biomarkers of neuroinflammation and progression to neurodegeneration in Alzheimer’s disease. J Neuroinflammation 2016;13:194. 10.1186/s12974-016-0670-z.

[41] Iannucci J, Grammas P. Thrombin, a Key Driver of Pathological Inflammation in the Brain. Cells 2023;12:1222. 10.3390/cells12091222.

[42] Lee R-L, Funk KE. Imaging blood–brain barrier disruption in neuroinflammation and Alzheimer’s disease. Front Aging Neurosci 2023;15:1144036. 10.3389/fnagi.2023.1144036.

[43] Guerreiro R, Wojtas A, Bras J, Carrasquillo M, Rogaeva E, Majounie E, et al. TREM2 Variants in Alzheimer’s Disease. New England Journal of Medicine 2013;368:117–27. 10.1056/NEJMoa1211851.

[44] Wang L, Nykänen N-P, Western D, Gorijala P, Timsina J, Li F, et al. Proteo-genomics of soluble TREM2 in cerebrospinal fluid provides novel insights and identifies novel modulators for Alzheimer’s disease. Mol Neurodegeneration 2024;19:1. 10.1186/s13024-023-00687-4.

[45] Kunkle BW, Grenier-Boley B, Sims R, Bis JC, Damotte V, Naj AC, et al. Genetic meta-analysis of diagnosed Alzheimer’s disease identifies new risk loci and implicates Aβ, tau, immunity and lipid processing. Nature Genetics 2019;51:414–30. 10.1038/s41588-019-0358-2.

[46] Zou K, Maeda T, Watanabe A, Liu J, Liu S, Oba R, et al. Aβ42-to-Aβ40-and Angiotensin-converting Activities in Different Domains of Angiotensin-converting Enzyme*. Journal of Biological Chemistry 2009;284:31914–20. 10.1074/jbc.M109.011437.

[47] Zou K, Yamaguchi H, Akatsu H, Sakamoto T, Ko M, Mizoguchi K, et al. Angiotensin-converting enzyme converts amyloid beta-protein 1-42 (Abeta(1-42)) to Abeta(1-40), and its inhibition enhances brain Abeta deposition. J Neurosci 2007;27:8628–35. 10.1523/JNEUROSCI.1549-07.2007.

